# Automated and objective analysis of speech in premanifest and early-stage Huntington’s disease

**DOI:** 10.1101/2022.10.27.22281625

**Authors:** Adam P. Vogel, Cheuk S. J. Chan, Geoffrey W. Stuart, Paul Maruff, Yenni Lie, Julie C. Stout

## Abstract

**Background:** Clinical markers that show change in performance in people with Huntington’s disease (HD) during the presymptomatic and prodromal stages remain a target of investigation in clinical medicine. Alongside genetic and neuroimaging initiatives, digital speech analytics has shown promise as a sensitive clinical marker of premanifest HD.

**Objective:** To investigate the sensitivity of digital speech measures for detecting subtle cognitive-linguistic and fine motor features in people carrying the expanded HD gene, with and without symptoms.

**Methods:** Speech data were acquired from 110 participants (55 people with the expanded HD gene including 16 presymptomatic HD; 16 prodromal HD; 14 early-stage HD; 9 mid-stage HD; and 55 matched healthy controls). Objective digital speech measures were derived from speech tasks that fit along a continuum of motor and cognitive complexity. Acoustic features quantified speakers’ articulatory agility, voice quality and speech-timing. Subjects also completed the tests of cognition and upper limb motor function.

**Results:** Some presymptomatic HD (furthest from disease onset) differed to healthy controls on timing measures derived from the syllable repetition and monologue. Prodromal HD presented with reduced articulatory agility, reduced speech rate and longer and variable pauses. Speech agility correlated with poorer performance on the upper limb motor test.

**Conclusion:** Tasks with a mix of cognitive and motor demands differentiated prodromal HD from their matched control groups. Motor speech tasks alone did not differentiate groups until participants were relatively closer to disease onset or symptomatic. Data demonstrated how ubiquitous behaviors like speech, when analyzed objectively, provide insight into disease related change.

## Introduction

Huntington’s disease (HD) is an autosomal dominantly inherited neurodegenerative disease caused by an unstable expansion in the number of CAG trinucleotide repeats in the 1T15 huntingtin (*HTT*) gene ^1, 2^. Its dominant inheritance means that prediagnostic genetic testing can be used to identify people at risk of developing HD prior to reaching diagnostic certainty ^3^. Investigation of both biological and symptomatic changes in people carrying the repeat expansion for *HTT* provides a basis for development of brain behavior models of early HD. During this stage, termed the premanifest stage ^4^, changes may be observed across cognitive, behavioral, fine motor and/or speech functions in HD CAG expansion, potentially years or decades before the emergence of diagnostic motor symptoms ^5-7^. A clinical diagnosis of HD is only made when a person carrying the expanded HD gene presents with unequivocal motor symptoms ^4^.

The genetic predictability of HD also provides an opportunity for introducing preventative or curative genetic and pharmaceutical therapies ^8^. One of the anticipated challenges of clinical trials in this premanifest stage is the need for sensitive and reliable markers of disease expression and response to treatment ^9-11^. Once people are symptomatic (manifest), HD impacts cognitive, linguistic, and motor function ^7, 12-15^. Qualitatively similar but quantitatively less severe changes in some of these symptoms have been reported in the prodromal period ^16-19^. There are, however, conflicting reports on the nature of speech changes occurring prior to an HD diagnosis ^7, 20^. Further, the relationship of fine motor function to speech in premanifest HD remains largely unknown.

Here we explored the use of speech analytics to provide continuous, quantitative data on disease status in HD. The aim was to investigate whether speech differed to healthy controls in premanifest (presymptomatic, prodromal) or manifest (early, mid) HD groups and to explore the relationship between these features and changes in fine motor control.

## Methods

### Participants

Fifty-five participants carrying the expanded huntingtin (HTT) gene and 55 age and sex matched controls were recruited from the ENROLL-HD registry at Calvary Health Care Bethlehem Hospital and a Monash University HD research volunteer database in Melbourne, Australia. All participants in the HD groups had ≥ 38 CAG repeats in one of the HD alleles. Participants received a standard neurological exam by a neurologist using the Unified Huntington’s Disease Rating Scale (UHDRS) yielding a Total Motor Score and a Diagnostic Confidence Level ^4, 21^. Potential HD participants were excluded if they presented with any comorbid neurological diseases that could affect performance on study measures (e.g., stroke, multiple sclerosis); clinical symptoms other than those caused by HD; a history of communication disorder (e.g., stuttering); lack premorbid competency in English; a history of alcohol or drug abuse that required medical intervention; or a history of learning disability and/or intellectual impairment. Eligibility for inclusion as a healthy control required having no family history of HD, or other major neurological disease, and had unremarkable cognition, speech, or language function. English was the first language for all participants.

Disease burden scores (DBS) were calculated for all participants with the expanded HD gene (DBS = age*[CAG repeat -35.5]) ^22^. Years to disease onset was estimated for participants with premanifest HD ^23^. Participants were allocated to one of four groups, based on proposed criteria from Ross et al., 2019: premanifest (presymptomatic, prodromal) or manifest (early, mid) groups (Table 1). Participants were included in the presymptomatic group if they were allocated a DCL of 0 or 1 and presented with an absence of motor and cognitive symptoms related to HD. Participants were allocated to the prodromal group if they presented with subtle motor signs, with or without minor/major neurocognitive changes, which corresponded with a DCL of 2 or 3 (Ross et al., 2019). Participants with a DCL of 4 were classified using the UHDRS Total Functional Capacity Rating Scale (TFC) scores as early HD (TFC ≥ 7) or mid-stage HD (TFC 4-6) ^24^. There were four control groups, each matched to the sex and age of the corresponding experimental group (see Table 2 for clinical and demographic information and Supplementary Materials).

**Table 1:** Participant group descriptions and inclusion criteria of HD groups.

|  | **HD groups** | **Criteria** | **Group description** |
| --- | --- | --- | --- |
| Premanifest phase | Presymptomatic HD | DCL = 0 to 1 | • No detectable cognitive changes<br>• Possible nonspecific minor motor findings that represent the individual's baseline |
|  | Prodromal HD | DCL = 2 or 3 | • Subtle motor decline from premorbid level of function, with or without minor or major neurocognitive changes |
| Manifest phase | Early HD | DCL = 4<br>TFC = 7 – 13 | • Clinical motor symptoms of HD<br>• Detectable change in TFC |
|  | Mid-stage HD | DCL = 4<br>TFC = 4 – 6 | • Clinical motor symptoms of HD<br>• Extensive detectable changes in TFC |

Note: DCL = diagnostic confidence level; TFC = Total Functional Capacity

**Table 2:** Demographic, clinical, and genetic characteristics of participants with the expanded HD gene (Mean +/-SD unless specified)

|  | **Presymptomatic**<br>**(n=16)** | **Prodromal**<br>**(n=16)** | **Early**<br>**(n=14)** | **Mid**<br>**(n=9)** |
| --- | --- | --- | --- | --- |
| Age<br>(years) | 41.1 (13.8) | 44.0 (10.6) | 56.1 (10.7) | 48.7 (13.7) |
| Sex<br>(female) | 12 (75%) | 10 (62.5%) | 2(14.3%) | 4 (44.4%) |
| DCL | 0.13 (0.34) | 1.56 (0.73) | 4.0 (0.0) | 4.0 (0.0) |
| DBS <sup>a</sup> | 240.0 (84.6) | 305.4 (94.2) | 418.7(116.9) | 362.9(129.6) |
| Range of<br>DBS | 94.5 – 364 | 143 – 480 | 324.5 – 714 | 150 – 532.5 |
| UHDRS-TMS <sup>b</sup> | 0.3 (1.0) | 4.3 (5.5) | 18.1 (7.6) | 32.7 (11.2) |
| TFC scores<br><sup>c</sup> | 13.0 (0.0) | 12.6 (1.0) | 10.0 (2.5) | 6.4 (1.5) |
| Range of<br>TFC | 13-13 | 10-13 | 3-13 | 4-9 |
| CAG<br>repeats | 41.8 (2.6) | 42.6 (2.1) | 43.1 (4.0) | 44.8 (5.3) |
| Estimated<br>YTO <sup>d</sup> | 18.8 (10.7) | 12.7 (8.1) | n/a | n/a |
| Disease<br>duration | n/a | n/a | 3.1 (1.5) | 6.3 (2.9) |

Note: DCL = diagnostic confidence level; DBS = disease burden scores; SD = standard deviation; UHDRS-TMS = The United Huntington's Disease Rating Scale – Total motor score; TFC = Total Functional Capacity; YTO = estimated years to disease onset; <sup>d</sup> Estimated years to disease onset was calculated using probability formulas <sup>23</sup>.

## Materials and Stimuli

We recorded speech in a quiet room using a standard laptop computer, a cardioid condenser (head-mounted) microphone (AKG520, Harman International Industries), coupled with an audio interface (Quad-Capture USB 2.0, Roland, Tokyo, Japan). Speech tasks included (i) sustained vowel /a:/, (ii) syllable repetition (DDK) (/papa/, /pata/), (iii) saying days of the week, (iv) reading a phonetically balanced passage from “The North Wind and The Sun”, and (v) an unprepared monologue for approximately one minute. Tasks varied along a spectrum of cognitive-motor complexity ^10, 25, 26^. All speech tasks except the monologue were repeated to minimize the impact of unfamiliarity ^27, 28^.

### Acoustic analysis of speech

Objective measures of timing and voice quality were derived from samples using acoustic analysis. Timing metrics relating to syllabic rate, silence length and their variability were derived from monologue, reading, days of the week and syllable repetition tasks as described previously ^10, 29^. Voice quality measures focused on (i) vocal fold vibration patterns, (ii) incomplete vocal fold closure, and (iii) placement of the articulators during vowel phonation were derived from the vowel ^30^. Detailed descriptions of each acoustic measure employed in this study are described in Supplementary materials Table S1.

### Cognitive and upper limb fine motor assessments

The Montreal Cognitive Assessment (MoCA) ^31^ and the Cogstate Brief Battery (CBB) were used to measure cognitive function. The MoCA is used as a cognitive screener with healthy controls scoring ≥ 26/30. The CBB battery assesses processing speed, attention/vigilance, working memory, and learning ^32^, and has demonstrated sensitivity in other neurodegenerative conditions ^33^. The Purdue pegboard test (PPT) was administered to all participants to assess fine motor performance (Model 32020, Lafayette Instrument Co., Lafayette, Indiana) ^34^. The PPT requires participants to insert metal pegs into a vertical row of holes using their dominant and non-dominant hands separately, proceeded by using both hands simultaneously and alternating hands.

### Statistical analysis

Statistical analyses were performed using Stata statistical software, version 17. All acoustic data extracted from premanifest, and manifest HD groups were compared to control data from this study. Due to violations of normality, with the HD group data distributions tending to have longer tails (see Figure 2 for examples), a nonparametric dominance statistic was used to compare groups ^35^. This statistic quantifies the probability that a randomly chosen participant from the HD group will have a worse score on a given measure than a randomly chosen participant from the control group (assessed across all possible pairs), with a value of 0.5 representing the null hypothesis. Significance tests used exact probability estimation. Confidence intervals were constructed using bootstrap resampling using bias-corrected and accelerated estimation ^36^ as recommended here ^37^. False discovery rate adjustment, assuming positive regression dependency, was used to adjust for multiple comparisons ^38^. To explore multivariate contrasts between groups, a small subset of measures that showed the best discrimination between HD and control groups was selected for further analysis. To avoid overfitting and to deal with correlations between the measures, elasticnet regression ^39^ was employed to construct composite scores for multivariate discrimination using logistic regression. Pearson correlation coefficients were also used to explore the relationships between clinical variables and speech outcomes.

### Multivariate Analysis

To evaluate how speech and motor measures might combine to discriminate HD gene carriers from controls in the pre-symptomatic and prodromal groups, we first selected key measures from the univariate analysis shown in Figure 1. This was to prevent overfitting when performing multivariate analysis, given the limited sample sizes. Two composite scores were developed using elasticnet logistic regression. The first included univariate measures of speech and motor for analysis, and the second included speech measures only. Motor performance, independent of speech, was assessed using Purdue pegboard right/left/both measures.

**Figure 1:**
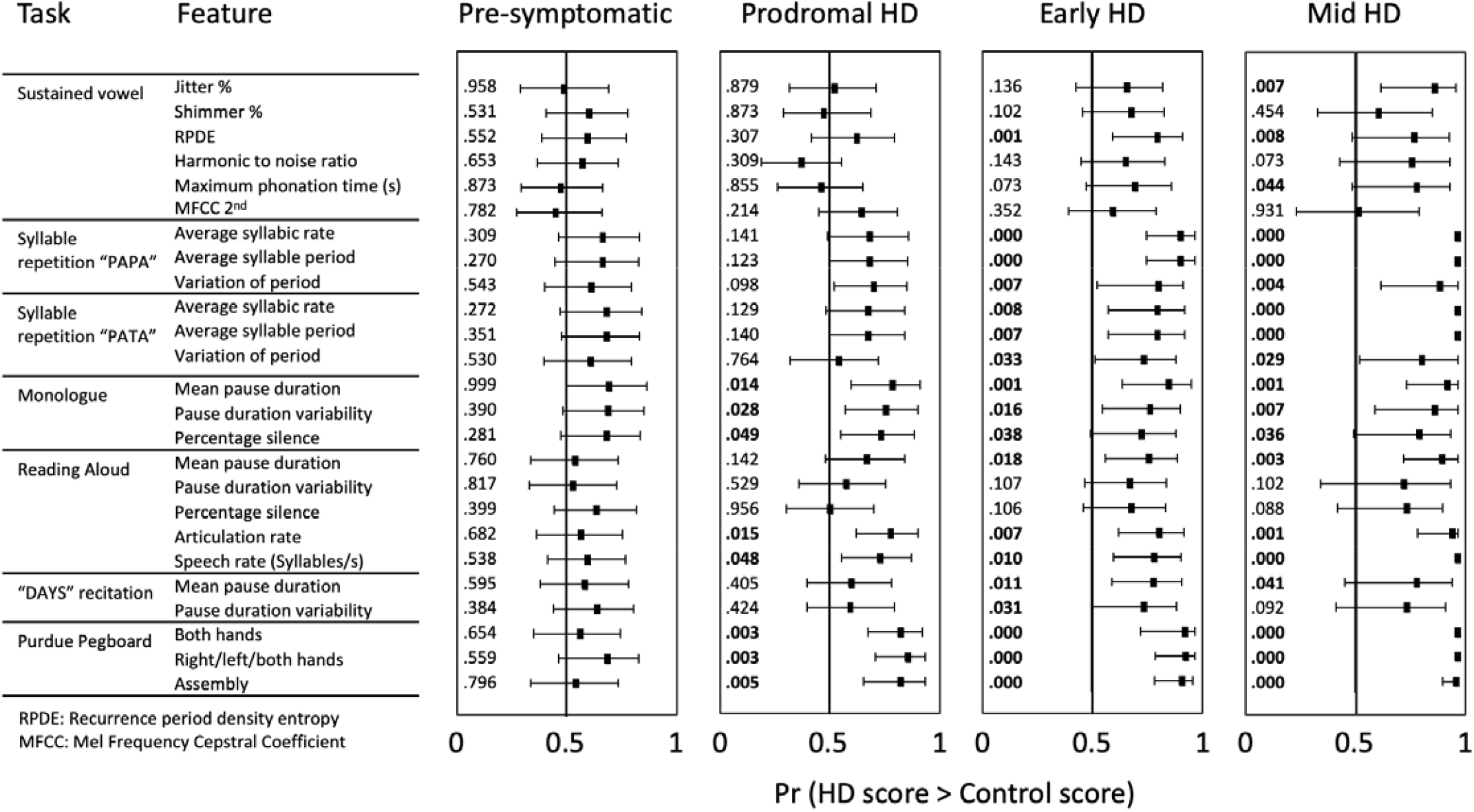
Standardized comparison of speech in premanifest and manifest HD to matched controls. Note: Scores of 0.5 (horizontal axis) indicate complete overlap of groups. Scores closer to 1 indicate separation of groups in the direction of increased deficit. Numerical values (q-values) in bold indicate tests with a false discovery rate <.05. Error bars represent 95% confidence intervals.

For the prodromal group, the elasticnet-derived composite score for speech measures was derived from the standardized scores from several speech tasks. The general composite score consisted of these same speech measures, plus performance on the Purdue pegboard test. These equally weighted composite scores were highly correlated with the exactly weighted scores from elasticnet regression (speech: r=0.976; general: r=.994). In the case of the prodromal group, the exact weights from elasticnet regression were far from equal, and so equal weights were not used. The speech composite score consisted of the measures derived from several tasks. The general composite score was derived from of the measures Purdue pegboard, and speech tasks. These composite measures were also highly correlated with exact regression scores (speech: r=989; general: r=.957).

## Results

Measures of speech timing and voice quality differed between HD participants and matched study controls, with the size of the effect increasing with disease severity (Figure 1). The largest differences between HD and controls were observed in manifest cohorts (premanifest < early < mid). Acoustic measures of timing derived from the monologue and syllable repetition tasks differentiated premanifest participants from matched controls. Voice quality was different between manifest and matched study participants, but not premanifest stages. As shown in Figure 2, the composite measures improved discrimination between all HD (premanifest and manifest) participants and matched study controls relative to the single measures.

### Speech in premanifest HD

No statistically significant differences (q-values <.05, representing the false discovery rate associated with each measure) were observed between presymptomatic and study control participants. Timing measures derived from the monologue and syllable repetition tasks revealed some presymptomatic participants did not fit within the healthy control range, appearing as outliers on several speech features (see Figure 2 and Table 3). The prodromal group differed to study controls on timing measures drawn from the monologue and reading tasks as well as for the Purdue motor task.

**Figure 2:**
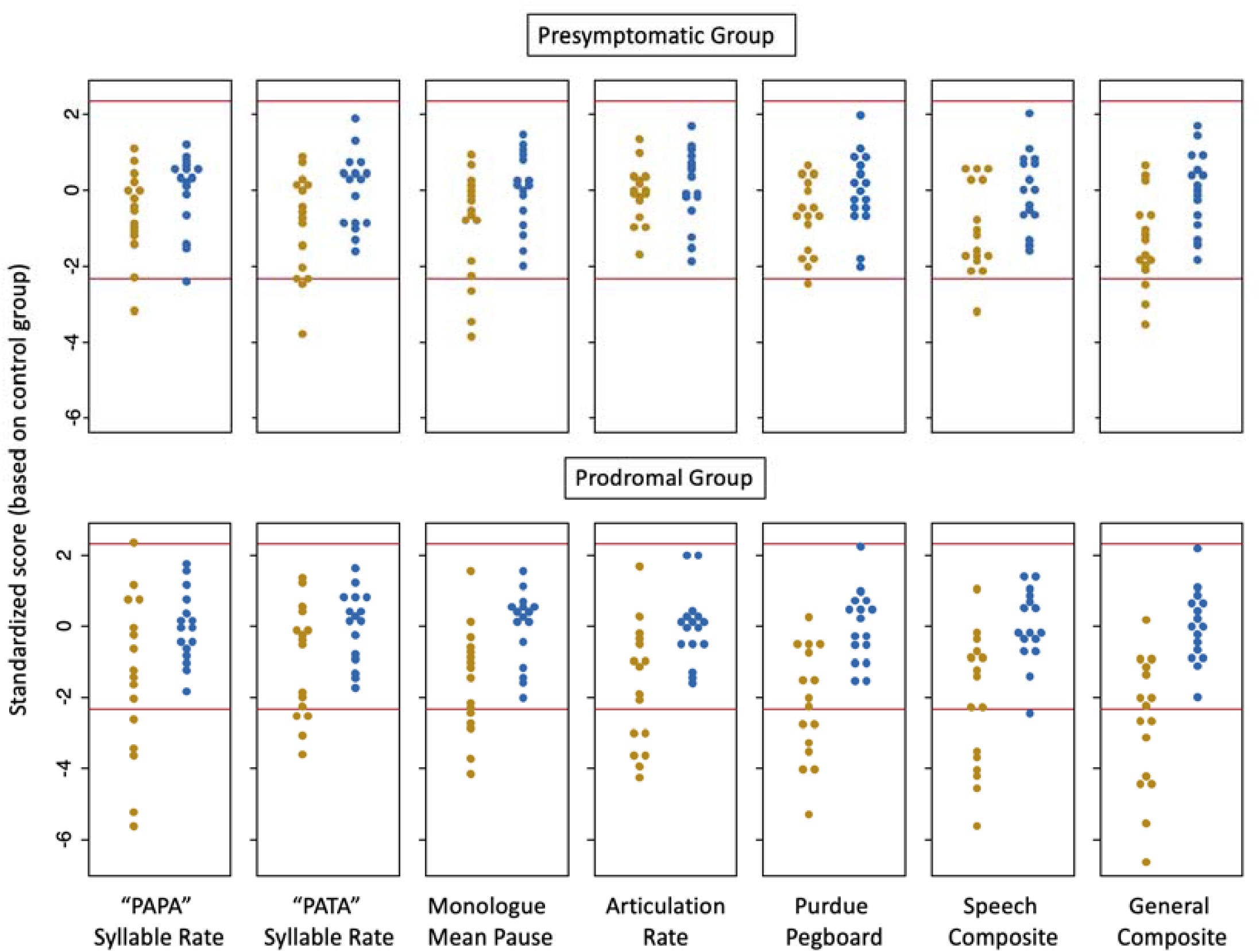
Standardized participant performance across core speech tasks compared to matched controls in presymptomatic and prodromal HD. Each dot represents a single participant. Red lines represent the upper and lower 1% confidence bounds of the control distributions (Z greater than 2.326). Participant scores outside of these boundaries means their performance sits beyond what is expected for age and sex matched healthy adults. All matched controls fit within the normal range and are represented by blue dots. HD participants are represented by yellow dots.

**Table 3:**
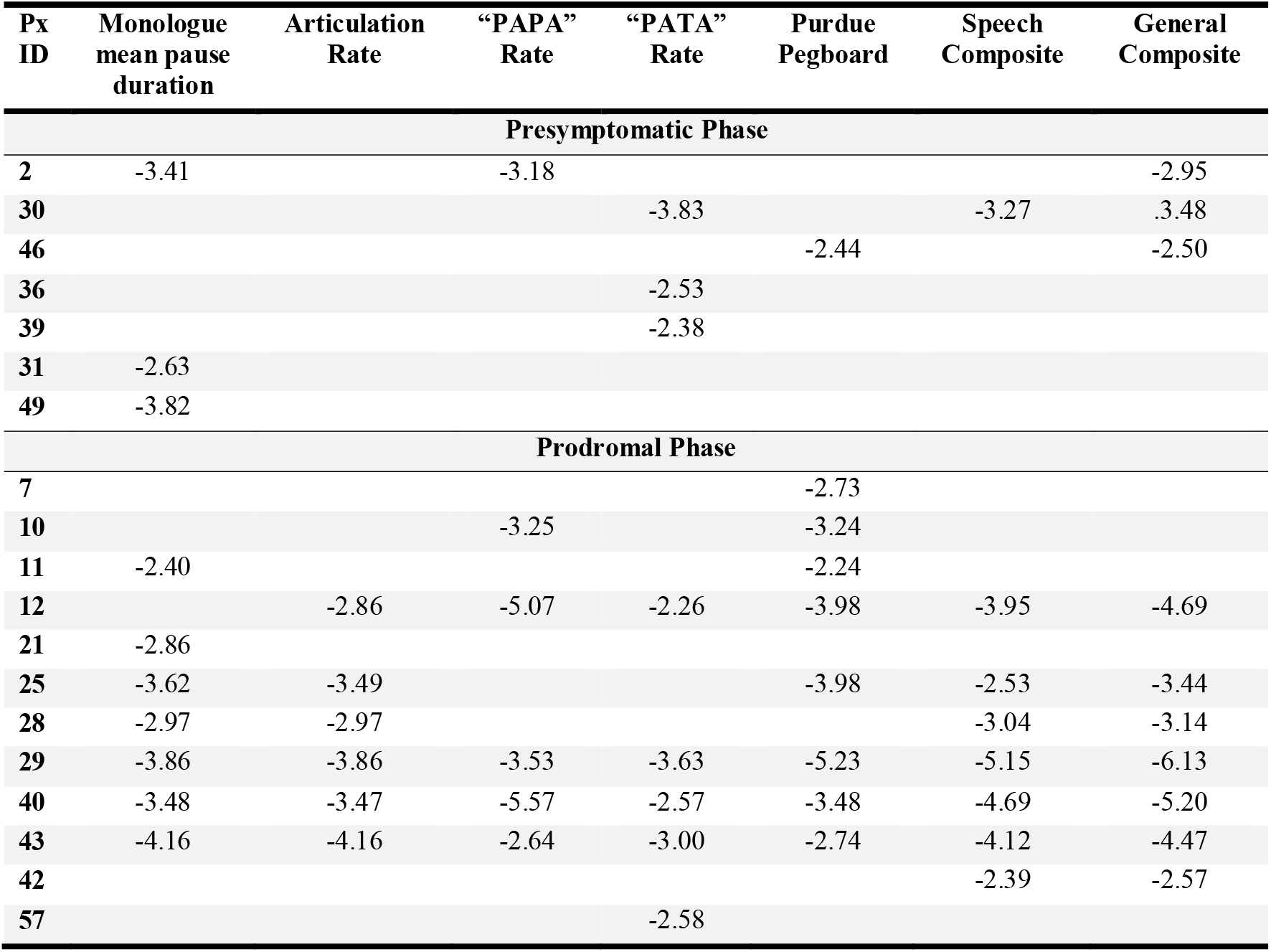
Premanifest HD CAG expansion carriers who are outliers on any selected measure, or on a composite score that includes the Purdue Pegboard test (General) or only speech measures (Speech).

### Speech in manifest HD

Speech in Early manifest HD was characterized by a slower and more inconsistent syllable repetition rate, increased irregularity in vocal fold vibration patterns (RPDE: q*=*0.001) during vowel phonation, and speech-timing deficits in complex speech tasks (reading: articulation rate: q = 0.01; percentage of silence: q=0.007 | monologue: mean pause duration: q=0.001; pause duration variability: q*=*0.028). Participants in the mid-stage manifest HD group presented with near global deficits across voice quality (jitter %: q=0.007), breath support (maximum phonation time: q=0.044) and measures of timing across complex tasks.

### Premanifest outlier performance across assessment domains

Figure 2 shows premanifest participants appearing as outliers relative to the control distributions across assessment domains. We examined whether these outlier participants were represented across different tests (Table 3). There appears to be a dissociation between the speech measures and the purely motor measure (Purdue). In the Presymptomatic group, four participants had deficits across individual speech measures and but not for the Purdue pegboard. One shows a specific deficit on the Purdue pegboard only. For the Prodromal group, most outliers presented with both speech and Purdue pegboard tasks. On participant (42) is a “multivariate outlier” (not an outlier on any individual measure) and two participants were outliers on individual speech measures but not the composite.

## Discussion

An automated speech analysis was used to separate premanifest and manifest HD from age and sex matched participants. Timing metrics derived from complex connected speech tasks identified healthy adults carrying the HD CAG expansion but not yet presenting with overt diagnostic signs. In some cases, abnormal speech timing was accompanied by pure motor deficits, with speech measures outperforming the pegboard, representing 85% of outlier instances in the presymptomatic group. Relative to matched healthy adults, participants in the premanifest groups demonstrated speech timing deficits during cognitively loaded tasks like reading and unprepared monologues, as well as the syllable repetition task, but not during recitation tasks like days of the week or sustained vowel tasks. The overall size of deficit relative to controls increased in the manifest groups, with the largest impact of dysarthria exhibited in the mid stage manifest HD group.

Two-thirds of presymptomatic HD participants produced speech at a similar rate to healthy adults, with the remaining third exhibiting longer pauses and slower syllabic rate than anticipated. A larger proportion of the prodromal group produced speech outside the normal range on at least one speech measure. Like presymptomatic HD, but to a larger extent, the prodromal HD group produced slower speech, with longer pauses across the monologue and syllable repetition tasks. While speech deficits in prodromal HD have been reported ^7, 16^-^18, 40^, our study is the first to successfully capture them in premanifest HD using an automated speech analytics pipeline.

The purely manual motor (Purdue) and speech measures both contributed important information to separating groups despite being moderately associated. At an individual level, several premanifest participants showed the emergence of HD motor signs in one or both measures suggesting that a single participant level analysis adds critical insights to the group level analyses. At a group level the shift in the set of scores was not uniform. Yet, some participants with the HD CAG expansion performed well outside the normal range in the presymptomatic and prodromal phases.

Speech measures such as articulation rate and mean pause duration on the monologue task best classified premanifest individuals from controls. The unprepared monologue is a complex communication task requiring rapid formulation of novel language and contemporaneous motor planning across multiple speech systems (i.e., respiration, phonation, prosody, resonance, articulation) ^25^. Unlike the recitation (days of the week) and syllable repetition tasks, which require relatively little linguistic planning, the monologue draws on cognitive domains like working memory, executive function, and lexical-semantic access. These additional demands on cognitive-linguistic performance perhaps contribute to the sensitivity of the monologue to early disease, similar to other cognitive/motor disorders like Fronto-temporal dementia ^26^ and spinocerebellar ataxia ^10^.

A composite of speech measures derived from the monologue and other tasks added to overall discriminability between groups. Like a listener’s ability to perceive an overall deficit based on a speaker’s communication in conversation, composite measures can encompass features across multiple speech subsystems, capturing performance beyond isolated and potentially esoteric acoustic features ^41, 42^. Unlike listener ratings however, digital acoustic analysis of speech is objective and is repeatable irrespective of clinical experience or training, making it better suited to monitoring change resulting from disease progression or treatment response ^28^.

Voice quality did not reliably separate HD groups from controls until individuals became symptomatic (i.e., early HD), similar to some earlier work ^43^ but contrary to other earlier reports ^17^. These conflicting outcomes are likely the result of different recording and analysis paradigms, or cohort characteristics of the premanifest HD population. Earlier studies would also not have differentiated between presymptomatic and prodromal stages when describing premanifest HD, given that this is new terminology in the HD field.

### Limitations and future work

Despite being the first study to explore speech in two tightly phenotyped premanifest groups alongside two symptomatic cohorts of HD, our sample size is modest. Replication and extension of our methodology could explore the use of home-based assessments and consortia-based data collection to increase cohort size and statistical power. The progression of HD and evolving speech phenotype across each disease stage suggests that composite measures need to be tailored to individual characteristics and severity. We have proposed a composite feature best suited to describing premanifest participants however these properties may change given that all speech systems are impacted in early to mid HD. Future research could also align structural and functional brain imaging, neurofilament light (NfL) concentrations ^44^, associated behavioral data (e.g., neuropsychological) and the role of medications (e.g., antipsychotic) with detailed speech outcomes to better describe the underlying mechanisms preceding or causing the HD dysarthria profile.

### Conclusion

Automated digital speech tests have potential to discriminate premanifest HD from controls at the single-person level. These changes appear to worsen as disease progresses. Digital markers that draw on multisystem behaviors (e.g., respiration, muscular control, cognitive-linguistic performance) to produce composite measures appear to outperform single feature metrics for differentiating disease group in HD. The ease of elicitation and administration of speech tests in a clinical trial setting compared to other modes of assessment (e.g., pegboard tests) highlights their potential for use beyond disease characterization.

## Data Availability

Data form part of consortia and ongoing initiatives

## Authors’ Roles

Design: APV, CSJC, JS

Execution: APV, CSJC

Analysis: APV, CSJC, GWS, YL, PM, JS

Writing: APV, CSJC

Editing of final version of the manuscript: APV, CSJC, GWS, YL, PM, JS

## Statistical Analysis

was conducted by Geoffrey W. Stuart PhD, Redenlab Inc, Victoria, Australia

