## Supplementary Table 1 for "Automated and objective analysis of speech in premanifest and early-stage Huntington’s disease"

Table S1
*Description of speech tasks and acoustic features.*

| Acoustic feature | Abbreviation | Description |
| --- | --- | --- |
| Syllable repetition (/pataka/, DDK) | | |
| Average DDK rate | DDK_avr | Average number of syllables produced (s) |
| Average DDK period | DDK_avp | Average period or duration between consonant-vowel syllable (CV) voicing offsets (ms) |
| Coefficient variation of DDK period | DDK_cvp | Variance of DDK period (%) |
| Breath support and Aerodynamic performance | | |
| Maximum phonation time | MPT | Maximum length of a continuous vowel phonation on one breath to indicate airflow sufficiency and adequacy of vocal folds closure (s) |
| Vocal fold vibration patterns – Aperiodicity and deviations in | | |
| Jitter | Jitt% | Cycle-to-cycle *f*0 variations ^1^ |
| Shimmer | Shim% | Fluctuations in amplitude |
| Recurrence period density entropy | RPDE | Deviations from periodicity in vocal fold vibration, or the uncertainty in estimating the duration of the vocal fold cycle ^2^ |
| Incomplete vocal fold closure – Aerodynamic vortices and acoustic noise | | |
| Harmonic-to-noise ratio | HNR | Estimates the level of noise in voice by comparing the periodic and non-periodic components of speech signals |
| Articulatory placements – Mel frequency cepstral coefficients (MFCCs) | | |
| 2^nd^ MFCC coefficient | 2^nd^ MFCC | A family of parameters that characterises fluctuations and postural instability of articulators during sustained vowel phonation |
| Prosody – Speech-timing parameters ^3-5^ | | |
| Mean pause duration | PMEAN | Average length of silence in a speech sample |
| Pause duration variability | PSTDEV | Variance of pause length |
| Percentage of silence | PPERC | Proportion of silence derived from total pause duration divided by total signal time |
| Rate of speech | SRATE | Speech rate derived from number of syllables per second |

**Additional information on group allocation using cognitive testing**

An absence of cognitive symptoms was established using the MoCA and Cogstate tests. Scores ≥26 on the MoCA were considered within normal limits ^6^. Most participants with DCLs of 3 or less scored above this threshold suggesting the MoCA was not sensitive to subtle cognitive deficits potentially manifesting in this group. Scores from four Cogstate cognitive tests were then compared to age matched controls to determine deviation from healthy performance. Participants were considered to have cognitive impairment if they scored ≥1.5 standard deviations from control means on 2 or more tests and allocated to the Prodromal group.
